# Long term Follow-up of the TREOCAPA (Prophylactic TREatment Of the duCtus Arteriosus in Preterm infants by Acetaminophen) Study (TREOCAPA-LT): protocol and statistical analysis plan

**DOI:** 10.64898/2026.09.30.26364424

**Authors:** Andrei Scott Morgan, Jean-Christophe Rozé, Moreno Ursino, Corinne Alberti, Victorine Desplebin, Mélanie Durox, Régis Hankard, Majda Hamrit, Samantha Johnson, Ruth Kemper, Yannick Marcon, Morgane Michel, Jennifer Zeitlin, Gilles Cambonie

## Abstract

**Introduction:** The TREOCAPA (Prophylactic TREatment Of the duCtus Arteriosus in Preterm infants by Acetaminophen) trial was a pan-European, multi-centre, phase II/III trial investigating the impact of prophylactic paracetamol (also known as acetaminophen) given during the first 5 days of life for children born before 29 weeks’ gestational age (GA) on survival to 36 weeks’ post-menstrual age without severe neonatal morbidities. However, due to time restrictions, long-term follow-up was not planned as part of the initial trial despite recommendations for neonatal trials to include endpoints measuring efficacy after discharge from the neonatal unit and on-going debate about the long-term effects of paracetamol in the neonatal period. This protocol describes a study to follow-up participants in the TREOCAPA trial at 2 years corrected age initiated after the trial’s onset.

**Methods and analysis:** The TREOCAPA Long Term (TREOCAPA-LT) study population consists of all children included in the TREOCAPA phase III trial in centres in countries participating in the follow-up study who are aged between 23.5 and 27.5 months corrected age during the study period. Families are contacted by the clinical teams and a parent or guardian is invited to complete an internet-based (online) questionnaire. The primary outcome is cognition, as measured by the non-verbal Cognitive Scale standard score from the Parent Report of Children’s Abilities-Revised (PARCA-R) questionnaire. Secondary outcomes are moderate to severe neurodevelopment, health service use and costs. Data will be analysed according to the study groups to which children were randomised in the original TREOCAPA RCT.

**Ethics and Dissemination:** The TREOCAPA-LT protocol was first approved by ethics’ committees in France (the study’s sponsor) and subsequently by all participating countries. Results from TREOCAPA-LT will be published in open access journals in accordance with the funder’s regulations.

**Registration:** https://clinicaltrials.gov/study/NCT06064825

**Strengths and limitations:**

- This follow-up study of a large multi-country randomised controlled trial of paracetamol use in preterm children born below 29 weeks of gestation is the first to examine long term effects of paracetamol without the influence of potential confounding factors in this vulnerable population.
- The study builds on an existing questionnaire in multiple European languages to assess cognition at 2 years of corrected age (PARCA-R) using newly developed questions to permit assessment of overall neurodevelopmental abilities as well as health economic analyses.
- Support in designing the study by the Global Foundation for Care of Newborn Infants (GFCNI) ensures that the study is relevant to patients and their families, and promotes knowledge sharing and public understanding of innovative research in neonatal care and preterm health.
- Despite differing regulatory and ethical requirements for participation between countries which caused some delays in study initiation, the number of eligible children is on target to achieve >80% power to detect a difference of 4.5 IQ points on a standardised scale – below the threshold of 6 which is considered clinically important.

## Introduction

The ductus arteriosus is a fetal connection between the pulmonary artery and aorta that diverts blood away from the lungs; persistence after initial transition to postnatal life can be associated with substantial mortality and morbidity in the neonatal period, and neonatal morbidities in turn may be associated with later deficits.[1] Non-steroidal anti-inflammatory drugs (NSAIDS – specifically, indomethacin and ibuprofen) have been used previously to facilitate ductal closure, but do not improve outcomes,[2–5] can have side effects[6] and are not recommended. [7–9] The most widely used analgesic drug in neonatal intensive care units in Europe after opioids is acetaminophen[10] (called ‘paracetamol’ in Europe and hereafter) which has been proposed as a safe alternative to NSAIDS for ductal closure.[11]

In this context, the TREOCAPA (Prophylactic **TRE**atment **O**f the du**C**tus **A**rteriosus in **P**reterm infants by **A**cetaminophen) trial was a pan-European, multi-centre, phase II/III trial (clinicaltrials.gov: NCT04459117) investigating the impact of prophylactic paracetamol given during the first 5 days of life for children born before 29 weeks’ gestational age (GA).[12,13] Phase II was a dose-finding study designed to identify the minimum effective dose of paracetamol for the closure of a patent ductus arteriosus (PDA) for neonates born at 23-26 weeks’ GA prior to inclusion of neonates born at these GAs in the phase III study.[14] Phase III was a pan-European, multi-centre, double-blind randomised controlled trial (RCT) with two arms in a 1:1 ratio examining the superiority of paracetamol to improve survival to 36 weeks of postmenstrual age (PMA) or first hospital discharge without severe neonatal morbidity by stimulating closure of a PDA compared to placebo in children born at 23-28 weeks’ GA inclusive; inclusions were stratified by gestational age group: 23-26 and 27-28 weeks’ GA.[12,13] Despite its ambition and scope (43 hospitals in 14 countries), long-term follow-up was not included in the TREOCAPA protocol due to time restrictions on the duration of the funding period, which was only sufficient to cover recruitment and follow-up during the initial hospitalisation until death or discharge from hospital. While neonatal endpoints are critical outcomes in themselves and often correlated with long-term health and development, they provide incomplete information, leading to calls for systematic follow-up of trials in very preterm populations.[15] Long term follow-up is needed to assess the potential longer term impact of improvements in neonatal clinical outcome as well as to rule out long-term adverse effects – i.e. to ensure that treatments are safe.

There is on-going debate about the long-term effects of paracetamol in the neonatal period.[16] Mice studies suggest exposure to paracetamol during neonatal brain development affects cognition.[17] In humans, a possible association between intra uterine neonatal exposure and later attention deficit, hyperactivity and autism has been suggested[18–20] but findings were susceptible to bias and may not be accurate.[18,21]

Given the ultimate aim of improving long-term outcomes through treatment of the patent ductus and the need to provide data regarding possible benefits or harms, we obtained additional funding to follow surviving children included in the trial up to two years’ corrected age.

### Research objectives and hypotheses

The objective of the TREOCAPA Long Term (TREOCAPA-LT) study is to assess outcomes of survivors at 2 years corrected age according to treatment group for surviving children included in the TREOCAPA trial. The primary hypothesis is that there will be improved cognitive development in children treated with paracetamol during the first 5 days of life.

Secondary objectives, related to effectiveness and safety, are, among children initially enrolled in TREOCAPA, to assess:

1. Survival without moderate or severe cognitive impairment at 2 years’ corrected age.
2. Survival without moderate or severe neurodevelopmental impairment at 2 years’ corrected age.
3. Health services use, including hospitalisations, and other care costs, and cost-effectiveness of paracetamol at 2 years’ corrected age.

The secondary hypotheses of the TREOCAPA-LT study related to these objectives are that, among children treated with paracetamol during the first 5 days of life, there will be:

1. Reduced moderate or severe neurodevelopmental impairment.
2. Reduced need for secondary hospitalisations following the initial neonatal hospitalisation.
3. Reduced long term health care utilisation costs associated with extremely preterm birth which, added to the increased effectiveness, will make for a cost-effective intervention.

## Methods and analysis

### Research design

TREOCAPA-LT is a multi-centre, pan-European observational study including children surviving to 2 years’ corrected age who were previously enrolled in the TREOCAPA phase III trial. Participation involves the parent or guardian completing an internet-based (online) questionnaire which is designed to take no longer than 20-30 minutes. This questionnaire is accessible via any web-connected device –- personal computer, tablet, or mobile (smart) telephone – with an alternative option to use paper questionnaires. No interventions are administered, nor is the child/family required to attend an in-person appointment, although it is possible for parents to complete the questionnaire during a routine follow-up visit, or for local investigators who were involved in the care of the child during the initial (neonatal) hospitalisation to obtain answers during a telephone call with the parents.

Data will be analysed according to the study groups to which children were randomised in the original TREOCAPA RCT –- intervention (prophylactic paracetamol during the first 5 days of life) or control (placebo). Local investigators, parents and the study team remain blinded to allocation.

In secondary analyses, the population included in the TREOCAPA trial will be compared with other European very preterm observational cohorts, allowing assessment of the generalisability/transportability of the trial results to other patient populations.

### Study population

The study population for TREOCAPA-LT consists of all children included in the TREOCAPA phase III trial in countries participating in the follow-up study. Given the complexity of setting up this study, countries were selected to participate in relation to the number of children recruited for the TREOCAPA trial and according to the availability of the PARCA-R questionnaire in national languages. Centres from 11 countries were initially invited to participate, but interpretations of regulatory procedures vary substantially across Europe which caused delays. In particular, Austria proved to be a particular outlier and, despite substantial time-consuming work to address regulatory obstacles and obtain ethical permissions, delays ran to over 18 months. When issues were finally resolved, study implementation was no longer logistically feasible in the remaining time frame of the project and, consequently, only 10 countries were included (Belgium, Denmark, Estonia, Finland, France, Greece, Ireland, Portugal, Sweden and Switzerland).

#### Inclusion criteria

- Included in the TREOCAPA phase III trial in participating centres;
- Aged between 23.5 and 27.5 months corrected age during the study period.

#### Non-inclusion criteria

The principal non-inclusion criteria are if:

- the local investigator does not have up-to-date contact information allowing contact with parents;
- the child’s vital status cannot be ascertained;
- the child is nearing the end of her/his life or experiencing a severe medical event as assessed by the local investigator;
- the child has become subject to a legal protection measure preventing their ongoing participation in clinical research;
- either parent or guardian opts out of participating (or does not provide consent in countries where it’s required);
- a language barrier prevents the parent or guardian from answering the questionnaires.

### Outcomes and assessment

#### Primary endpoint

The primary outcome is the Non-Verbal Cognitive (NVC) Scale standard score from the Parent Report of Children’s Abilities-Revised (PARCA-R) questionnaire[22] at 23.5 to 27.5 months’ corrected age.

#### Secondary endpoints

Secondary endpoints, assessed using the parental questionnaire, include:

1. Survival without moderate or severe cognitive impairment at 2 years corrected age, defined as a PARCA-R NVC scale score more than 2 standard deviations (SD) below the normative mean (score <70).
2. Survival without moderate or severe neurodevelopmental impairment based on the presence of at least one of the impairments below:

a. PARCA-R NVC scale score more than 2 SD below the normative mean (score <70);
b. moderate to severe gross motor impairment;
c. unilateral or bilateral deafness or blindness;
3. Health services use (as reported by parents), costs, and incremental cost-effectiveness ratio.

#### Parental questionnaire

The PARCA-R is a parent-report questionnaire,[22] also used in other RCTs (e.g. DOLFIN,[23] SAFEBoosC-III,[24] Baby-OSCAR,[25] SIFT,[26] OPTIMIST-A[27]), that is validated and age-standardised at 23.5 to 27.5 months of age, from which norm-referenced standard scores are derived with a normative mean of 100 (SD 15). The NVC Scale includes 34 items translated into languages covered by TREOCAPA-LT. The questionnaire can be completed by any adult who is a parent or carer of the child and is free available for use (see: https://www.parca-r.info).

Additional questions were developed based on professional society recommendations and questions used in previous studies -– notably, the EPICE/SHIPS preterm-birth cohort (11 European countries, 2011),[28] and the Baby-OSCAR trial[5,25,29] (ibuprofen treatment for a confirmed ductus arteriosus, UK). Questions cover the children’s general health status as well as health service use, including hospitalisations and time taken off from work by parents because of their children’s health needs. The whole study questionnaire is designed to take no more than 20-30 minutes per child to complete.

### Sample size

The expected final sample size is based on the number of inclusions in the TREOCAPA RCT, the countries participating in TREOCAPA-LT, mortality before age 2 and expected loss to follow-up or refusals to participate. We estimated this would be 400-500 children (200 to 250 by arm, i.e. 794 [number of inclusions targeted for TREOCAPA] x 0.8 [survival rate] x (0.7 or 0.85 [low and high follow-up rates]) – 40 [over the limit age of 27.5 months corrected for evaluation by the PARCA-R when the study commenced]). After the trial’s completion, the total number of inclusions in the main analysis was 778, with survival to hospital discharge of 86% (669) in TREOCAPA. However, 87 children were over evaluation age at commencement of TREOCAPA-LT because of longer than anticipated times for completing ethical, regulatory and logistic requirements in participating countries (see Table 1 for the start dates and numbers eligible at the study’s onset in each country) and not all countries or centres were included. This means a follow-up rate of at least 76% is required to obtain a minimum of 400 participants.

**Table 1:** Number of children and centres included in the TREOCAPA phase 3 study, by country.

| **Country** | **Number of centres in the TRECOPA trial (number included in TRECOPA-LT)** | **Number of children included in TRECOPA surviving to discharge** | **TRECOPA-LT study start date<sup>1</sup>** | **Number of children over the PARCA-R cut-off age and thus ineligible for inclusion in TRECOPA-LT** | **Number of children considered for inclusion in TRECOPA-LT<sup>2</sup>** |
| --- | --- | --- | --- | --- | --- |
| Austria | 1 (0) | 15 | — | — | — |
| Belgium | 1 (1) | 11 | 13 June 2024 | 1 | 10 |
| Denmark | 1 (1) | 11 | 17 June 2024 | 1 | 10 |
| Estonia | 3 (3) | 30 | 07 September 2024 | 3 | 27 |
| Finland | 5 (3) | 50 | 28 May 2025 | 15 | 20 |
| France | 16 (16) | 366 | 08 December 2023 | 10 | 353 |
| Greece | 3 (3) | 26 | 02 June 2025 | 9 | 16 |
| Ireland | 1 (1) | 15 | 22 November 2024 | 2 | 13 |
| Italy | 3 (0) | 7 | — | — | — |
| Norway | 1 (0) | 7 | — | — | — |
| Portugal | 4 (3) | 26 | 21 February 2025 | 13 | 13 |
| Spain | 3 (0) | 9 | — | — | — |
| Sweden | 1 (1) | 14 | 15 April 2025 | 4 | 10 |
| Switzerland | 3 (3) | 80 | 12 November 2024 | 29<sup>3</sup> | 51 |

1. For countries with multiple centres, the date corresponds to when the first centre opened.
2. Some children were excluded after vital status was ascertained.
3. Routine follow-up at one of the Swiss centres includes completion of the PARCA-R, hence data were available for the primary outcome for an additional 9 children.

We computed the power for Cohen’s d,[30] meaning the difference between the means of the primary outcome (a continuous score) divided by the pooled standard deviation, under different sample size scenarios for a fixed 2-sided type I error of 0.05. As shown in Table 2, with 200 children in each group, we will have at least 80% power, a Cohen’s d of 0.3, equivalent to 4.5 points on a standardised cognitive (PARCA-R/IQ) test – below the threshold of 6 which is considered clinically important for public health interventions.[31,32]

**Table 2:** Statistical power (%)of the TREOCAPA-LT study according to Cohen’s d and sample size in each arm of the study.

| **Cohen's d** | **Sample size by group (n)** |  |  |  |
| --- | --- | --- | --- | --- |
|  | **150** | **200** | **250** | **300** |
| **0.4** | 93.2 | 97.9 | 99.4 | 99.8 |
| **0.3** | 73.6 | 84.9 | 91.7 | 95.6 |

### Recruitment and selection

At hospital discharge, contact information was retained locally by clinical services along with the corresponding TREOCAPA ID. No contact information is available to the central research coordinators. Families are invited to participate by local investigators when the child is 23.5 months’ corrected age after ascertainment of the child’s vital status to avoid inappropriate contacts. Inability to ascertain vital status is a non-inclusion criterion for the TREOCAPA-LT study.

A list of participant IDs for each recruiting centre and the date at which the child turns 23.5 months’ corrected age is used by the central investigating team of TREOCAPA-LT to send a reminder to the local investigator to contact parents. This date is based on the child’s GA at birth and birth date, but the child’s age or GA at birth cannot be determined from this information alone, thus preserving patient-identifiable data from being disclosed. This information is used to send a reminder to the local investigator when the family should be contacted and to inform the investigator whether the family has opted out, filled in the questionnaire or needs a follow-up call (see below). This list is updated during the study as more children are recruited or eligibility status changes.

When the child turns 23.5 months’ corrected age (and the investigator is emailed), contact will be made by telephone from the local investigator to the parents (or legal guardian) of the child to provide information about the study, to invite them to participate and to collect the contact details (emails) of both parents if not already known. During this call, the investigator will ascertain which parent is the child’s main carer who will respond to the questionnaire. Parental contact information will only be used for sending information documents and the questionnaire. If only the address of the parents is available, a paper Information Notice and letter will be sent asking them to contact the local investigator.

If the parents or guardians are interested in participating, the investigator will send both parents a link to a detailed information leaflet about the study. For the person who will respond to the questionnaire, a personalised link is also sent with an invitation to connect to the online data collection tool. This tool first provides access to the detailed information document, and then to the questionnaire.

Alongside information about the study in the detailed information leaflet, the responding parent is given the option to either proceed to the questionnaire or to opt out of the study. The co-parent who is not responding has the option of emailing or telephoning the local investigator to opt out. The questionnaire can be consulted on the study website: https://treocapa.inserm.fr/. If either parent opts out of participating in the follow-up, no further reminders to participate will be provided.

Local investigators are notified by email when families have responded to the questionnaire. Reminders are sent to investigators if families fail to respond to the questionnaire so they can contact the parents again. Reasons why families were not contacted or why parents chose not to participate are noted by local investigators and provided to the coordination team.

### Data Collection

The OBiBa/Amber (https://amberdoc.obiba.org) questionnaire management software is an open-source application developed within the OBiBa software suite (https://www.obiba.org) and is designed for the secure and structured collection of individual-level data in epidemiological and public health studies.[33] For the TREOCAPA-LT study, Amber was extended with the following features: per-centre participant management, including eligibility tracking, the sending of reminders, and the generation of personal access tokens; interview design composed of multiple interdependent questionnaires; support for country-specific web interfaces with regional language variations; an API to access participant records for downstream analysis; and the inclusion of metrics to follow-up on study records.

### Statistical analysis plan

Investigators are blinded to the TREOCAPA study groups and will be throughout the follow-up period. Missing follow-up data (e.g. from non-responders) will be assessed to detect if missingness is due to an underlying mechanism and hence not at random. Imputation for PARCA-R scores for questionnaires returned after 27.5 months will be considered. Subgroup analyses will be performed by sex and gestational age (23-26 and 27-28 week groups) as per the original trial; no interim analyses are planned.

Unless otherwise specified, statistical significance is defined as p<0.05. Multiplicity adjustment will be considered.

### Analysis of the primary endpoint

The primary endpoint will be analysed according to the principle of intention to treat (ITT, i.e. analysis of all patients meeting the inclusion criteria for TREOCAPA-LT regardless of the treatment received in the initial TREOCAPA trial) after cessation of inclusions. Arm comparisons will be conducted using the parametric test for mean difference.

### Analysis of the secondary endpoints

Secondary analyses will be done both on an ITT and a “per-protocol” basis. Quantitative variables will be described as medians [quartiles] or means [standard deviation] depending on their distribution. Qualitative variables will be described as numbers [percentages]. Inter-group comparisons will be conducted using parametric or non-parametric tests according to the nature and the distributions of the variables.

Secondary criteria relating to health economic analyses will use an all-payers perspective for the main analysis; both a statutory health insurance perspective (to assess cost-effectiveness for the principal payer) and a societal perspective (to account for parents’ productivity losses) will be carried out. The time horizon will be 2 years. The effectiveness criterion will be life years gained; however, if there is no difference between the two groups in terms of survival, the effectiveness criterion will be survival without moderate or severe cognitive impairment. Costs will include direct medical costs (hospitalisations, consultations, medications) and non-medical costs (informal care). In the societal analysis, parents’ indirect costs (productivity losses) will be included. An incremental cost-effectiveness ratio will be calculated to determine the incremental costs needed to gain an additional life-year (or an additional life-year without moderate or severe cognitive impairment) with paracetamol compared to usual care. Both deterministic and probabilistic sensitivity analyses will be carried out to assess the uncertainty surrounding our results.

As it is not feasible to carry out a cost-effectiveness analysis in each participating country, resources consumed by children will be pooled and then valued using French unit costs (as France is the principal recruiter) as a proof of concept. Economic evaluation will be carried out using French guidelines.[34]

### Patient Involvement

The Global Foundation for Care of Newborn Infants (GFCNI, formerly the European Foundation for Care of Newborn Infants) played a key role in the original TREOCAPA trial[13] and continues its involvement in the TREOCAPA-LT study. GFCNI, as a representative of patients and parents, has been involved since the inception of the project and plays a key role to ensure that families’ voices remain central throughout the research process. In particular, GFCNI helped to develop and test the questionnaire, and works to raise awareness amongst affected families using educational materials to communicate the importance and outcomes of the study to a broader audience (https://www.gfcni.org/research/liftup). Efforts such as the production of a fact sheet about the importance of long term follow-up after clinical trials (https://www.gfcni.org/fileadmin/www.gfcni.org/Research/LIFT-UP_Preterm/ GFCNI_Factsheet_Clinical_Trials_in_Newborns_and_Children_EN_web.pdf) promote knowledge sharing and enhance public understanding of innovative research in neonatal care and preterm health.

### Ethics and dissemination

TREOCAPA-LT was designed to be conducted on a non-opposition basis, i.e. that consent for use of submitted data was implicit in participation after provision of a written notice with information about the study by the local investigator and being given the option to opt-out. However, requirements varied by country, with some requiring explicit consent; there was also variation in whether one or two parents needed to be informed and/or provide consent. Details by country, including relevant ethical approvals, are shown in Table 3.

**Table 3:** Ethical approvals for the TREOCAPA-LT study.

| **Country** | **Consent details (including whether non-opposition or explicit consent, and number of parents needing to be informed)\*** | **Ethical Board** | **Approval date** | **Registration number** |
| --- | --- | --- | --- | --- |
| Belgium | Explicit consent requiring form to be signed jointly by both parents. | Comité d’Ethique Hospitalo-Facultaire Universitaire de Liège | 11 April 2024 | 2024/142 |
| Denmark | Explicit consent signed by the parent/legal guardian attending the consultation. | National Scientific Ethics Committee | 06 February 2024 | Project ID: CVK-2009864; Notification number: 106599 |
| Estonia | Non-opposition | Research Ethics Committee of the University of Tartu (UT REC) | 20 May 2024 | Approval: 390/T-29 |
| Finland | Explicit consent signed by one parent/legal guardian only. | Regional Medical Research Ethics Committee of Northern Ostrobothnia | 20 February 2025 | PPHVA / 535/00.01.03.05/2022 |
| France | Non-opposition, but both parents need to be informed about the study | Committee for the Protection of Persons East | 04 September 2023 | SI number: 23.02385.000215<br>National number: 2023-A01418-37 |
| Greece | Non-opposition | Research Ethics and Conduct Committee of Aristotle University of Thessaloniki | 22 January 2025 | Meeting no. 134/22-01-2025 |
| Ireland | Consent signed by one parent/legal guardian only | Clinical Research Ethics Committee of the Cork Teaching Hospitals | 13 Aug 2024 | CREC Review Reference Number: ECM 4 (p) 30/07/2024 & ECM 5 (5) 02/07/2024 |
| Portugal | Consent form signed jointly by both parents/legal guardians | Ethic Committee of Santo Antonio, Porto | 5 June 2025 | N / RÉF. 2024.108(093-DEFI/095-CE) |
|  |  | Ethics Committee of Santa Maria, Lisbon | 22 January 2025 | RÉF. N° 247/24 |
|  |  | Ethics Committee of Hôpital São Francisco Xavier, Lisbon | 15 September 2025 | 2025-107 |
| Sweden | Consent signed by one parent/legal guardian, but both parents must have the information | Stockholm Ethic Committee | 02 April 2025 | 2024-08672-01 |
| Switzerland | Non-opposition | Cantonal Ethics Commission of Zurich | 25 October 2024 | 2024-01659 |

\*In all cases, only one parent completes the questionnaire.

Publications arising from the TREOCAPA-LT project will be published as open access in accordance with the regulations of the project’s financing. Should this not be possible, works will be published in an open access archive in accordance with Article 30 of the French *Loi pour une République Numérique*.[35] Further dissemination of results will be carried out in conjunction with GFCNI.

### Registration

https://clinicaltrials.gov/study/NCT06064825

## Discussion

This paper reports the study protocol for the TREOCAPA-LT study, a long-term observational follow-up at 2 years of age of children originally enrolled in the TREOCAPA randomised controlled trial.[13] Although funding wasn’t available for longer term follow-up as part of the original trial, planning for the 2 year follow-up commenced early and funding was obtained that allowed participant enrolment to start by mid-way through the recruitment period of the initial trial. This unfortunately meant some of the first children included in TREOCAPA were not eligible for the follow-up study as they were too old to participate by the time TREOCAPA-LT recruitment started. Furthermore, regulatory and ethical requirements – including interpretations of the European Union’s General Data Protection Regulation – varied substantially across recruiting countries. This caused substantial delays in a number of countries that impacted recruitment and, in one case, permissions arrived so late that no children could be included from that country. Statistical power will therefore be lower than it might have been had we been able to offer participation to more of the original TREOCAPA participants, but we remain on target for the minimum number of children required. Our experiences reflect the current difficulties of conducting pan-European research, poignantly illustrating the high regulatory burden and negative impact of varying interpretations of European regulations on research, and highlighting the importance of including follow-up in initial trial protocols.

The hypotheses for the TREOCAPA-LT follow-up study were formulated before the TREOCAPA trial results were known, hence were predicated upon the main hypothesis of the trial: that prophylactic paracetamol treatment during the first 5 days of life would improve short-term survival and decrease severe morbidities at 36 weeks’ PMA. We anticipated that the expected improved status at hospital discharge in the intervention group would result in improved cognitive development at 2 years of age. Given that no differences were seen between the treatment and control groups in the TREOCAPA trial,[13] it might now be considered unlikely that improved cognition will be seen at 2 years of age, similar to other recent reports.[29] However, follow-up remains important, particularly for addressing long-term safety concerns related to paracetamol, as the randomised design of the original trial ensures the potential influence of confounding factors is minimised.

## Conclusion

The pan-European TREOCAPA-LT observational follow-up study will permit the longer term effects of paracetamol administered to extremely preterm born children during the first few days of life to be examined.

## Data Availability

Not applicable (no data referred to in this manuscript)

## Authors’ contributions

ASM conceived and designed the study with support from GC, MU and JZ. Development of the original research protocol was led by ASM with contributions from GC, MU, CA, RH, SJ, RK, YM, MM, JCR and JZ. The questionnaire was based on the non-verbal cognitive (NVC) scale of the PARCA-R (Parent Report of Children’s Abilities-Revised) questionnaire (developed by SJ) with additional questionnaire sections developed by ASM, SJ, RK, MM and JZ. The online data collection software was developed by YM, and the questionnaire was implemented and managed by YM and MH. VD and MD were responsible for supporting ethics applications, managing the study and supporting the recruitment. ASM drafted the original manuscript. All authors critically revised the manuscript for important intellectual content. All authors approved the final manuscript.

## Acknowledgements

We would like to express our gratitude to the following people without whom it would not have been possible to implement TREOCAPA-LT: Aure Vanhecke – for assistance with the original funding submission; Gonçalo Gonçalves and Artur Rocha – for support with technical aspects; Barthélémy Tosello – for coordinating related projects; Alexandre Hoctin – for administrative help during recruitment; and the study sponsor: the French national institute for health and medical research (INSERM, Institute National pour la Sante et la Recherche Médicale).

## References

1 Chock VY, Bhombal S, Variane GFT, et al. Ductus arteriosus and the preterm brain. Archives of Disease in Childhood Fetal and Neonatal Edition. 2023;108:96–101. doi: 10.1136/archdischild-2022-324111

2 Schmidt B, Davis P, Moddemann D, et al. Long-term effects of indomethacin prophylaxis in extremely-low-birth-weight infants. The New England Journal of Medicine. 2001;344:1966–72. doi: 10.1056/NEJM200106283442602

3 Van Overmeire B, Allegaert K, Casaer A, et al. Prophylactic ibuprofen in premature infants: A multicentre, randomised, double-blind, placebo-controlled trial. Lancet (London, England). 2004;364:1945–9. doi: 10.1016/S0140-6736(04)17477-1

4 Gournay V, Roze JC, Kuster A, et al. Prophylactic ibuprofen versus placebo in very premature infants: A randomised, double-blind, placebo-controlled trial. Lancet (London, England). 2004;364:1939–44. doi: 10.1016/S0140-6736(04)17476-X

5 Gupta S, Subhedar NV, Bell JL, et al. Trial of selective early treatment of patent ductus arteriosus with ibuprofen. The New England Journal of Medicine. 2024;390:314–25. doi: 10.1056/NEJMoa2305582

6 Gournay V, Savagner C, Thiriez G, et al. Pulmonary hypertension after ibuprofen prophylaxis in very preterm infants. Lancet (London, England). 2002;359:1486–8. doi: 10.1016/S0140-6736(02)08424-6

7 Fowlie PW, Davis PG, McGuire W. Prophylactic intravenous indomethacin for preventing mortality and morbidity in preterm infants. The Cochrane Database of Systematic Reviews. 2010;Cd000174. doi: 10.1002/14651858.CD000174.pub2

8 Ohlsson A, Walia R, Shah SS. Ibuprofen for the treatment of patent ductus arteriosus in preterm or low birth weight (or both) infants. The Cochrane Database of Systematic Reviews. 2018;9:Cd003481. doi: 10.1002/14651858.CD003481.pub7

9 Mitra S, Scrivens A, Kursell AM von, et al. Early treatment versus expectant management of hemodynamically significant patent ductus arteriosus for preterm infants. The Cochrane Database of Systematic Reviews. 2020;12:Cd013278. doi: 10.1002/14651858.CD013278.pub2

10 Carbajal R, Eriksson M, Courtois E, et al. Sedation and analgesia practices in neonatal intensive care units (EUROPAIN): Results from a prospective cohort study. The Lancet Respiratory Medicine. 2015;3:796–812. doi: 10.1016/S2213-2600(15)00331-8

11 Allegaert K, Anderson B, Simons S, et al. Paracetamol to induce ductus arteriosus closure: Is it valid? Archives of Disease in Childhood. 2013;98:462–6. doi: 10.1136/archdischild-2013-303688

12 Ursino M, Alberti C, Cambonie G, et al. TREOCAPA: Prophylactic treatment of the ductus arteriosus in preterm infants by acetaminophen-statistical analysis plan for the randomized phase III group sequential trial. Trials. 2025;26:52. doi: 10.1186/s13063-025-08751-8

13 Roze J-C, Cambonie G, Flamant C, et al. Prophylactic treatment of patent ductus arteriosus with acetaminophen: A randomized clinical trial. JAMA Pediatrics. 2026;180:374–83. doi: 10.1001/jamapediatrics.2025.6150

14 Bouazza N, Cambonie G, Flamant C, et al. Prophylactic intravenous acetaminophen in extremely premature infants: Minimum effective dose research by bayesian approach. Paediatric Drugs. 2024;26:83–93. doi: 10.1007/s40272-023-00602-w

15 Marlow N, Doyle LW, Anderson P, et al. Assessment of long-term neurodevelopmental outcome following trials of medicinal products in newborn infants. Pediatric Research. 2019;86:567–72. doi: 10.1038/s41390-019-0526-1

16 Cendejas-Hernandez J, Sarafian JT, Lawton VG, et al. Paracetamol (acetaminophen) use in infants and children was never shown to be safe for neurodevelopment: A systematic review with citation tracking. European Journal of Pediatrics. 2022;181:1835–57. doi: 10.1007/s00431-022-04407-w

17 Viberg H, Eriksson P, Gordh T, et al. Paracetamol (acetaminophen) administration during neonatal brain development affects cognitive function and alters its analgesic and anxiolytic response in adult male mice. Toxicological Sciences : an Official Journal of the Society of Toxicology. 2014;138:139–47. doi: 10.1093/toxsci/kft329

18 Masarwa R, Levine H, Gorelik E, et al. Prenatal exposure to acetaminophen and risk for attention deficit hyperactivity disorder and autistic spectrum disorder: A systematic review, meta-analysis, and meta-regression analysis of cohort studies. American Journal of Epidemiology. 2018;187:1817–27. doi: 10.1093/aje/kwy086

19 Baker BH, Lugo-Candelas C, Wu H, et al. Association of prenatal acetaminophen exposure measured in meconium with risk of attention-deficit/hyperactivity disorder mediated by frontoparietal network brain connectivity. JAMA Pediatrics. 2020;174:1073–81. doi: 10.1001/jamapediatrics.2020.3080

20 Inoue K, Ritz B, Ernst A, et al. Behavioral problems at age 11 years after prenatal and postnatal exposure to acetaminophen: Parent-reported and self-reported outcomes. American Journal of Epidemiology. 2021;190:1009–20. doi: 10.1093/aje/kwaa257

21 Ji Y, Azuine RE, Zhang Y, et al. Association of cord plasma biomarkers of in utero acetaminophen exposure with risk of attention-deficit/hyperactivity disorder and autism spectrum disorder in childhood. JAMA Psychiatry. 2020;77:180–9. doi: 10.1001/jamapsychiatry.2019.3259

22 Johnson S, Bountziouka V, Brocklehurst P, et al. Standardisation of the parent report of children’s abilities-revised (PARCA-r): A norm-referenced assessment of cognitive and language development at age 2 years. The Lancet Child & Adolescent Health. 2019;3:705–12. doi: 10.1016/S2352-4642(19)30189-0

23 Parr J, Andrew M. Developmental outcomes of long-term feed supplementation in neonates – the DOLFIN randomised controlled trial (version 6.0). National Perinatal Epidemiology Unit CTU 2024.

24 Hansen ML, Pellicer A, Hyttel-Sorensen S, et al. Cerebral oximetry monitoring in extremely preterm infants. The New England Journal of Medicine. 2023;388:1501–11. doi: 10.1056/NEJMoa2207554

25 Gupta S, Juszczak E, Hardy P, et al. Study protocol: Baby-OSCAR trial: Outcome after selective early treatment for closure of patent ductus ARteriosus in preterm babies, a multicentre, masked, randomised placebo-controlled parallel group trial. BMC Pediatrics. 2021;21:100. doi: 10.1186/s12887-021-02558-7

26 Dorling J, Abbott J, Berrington J, et al. Controlled trial of two incremental milk-feeding rates in preterm infants. The New England Journal of Medicine. 2019;381:1434–43. doi: 10.1056/NEJMoa1816654

27 Dargaville PA, Kamlin COF, Orsini F, et al. Two-year outcomes after minimally invasive surfactant therapy in preterm infants: Follow-up of the OPTIMIST-a randomized clinical trial. Jama. 2023;330:1054–63. doi: 10.1001/jama.2023.15694

28 Zeitlin J, Maier RF, Cuttini M, et al. Cohort profile: Effective perinatal intensive care in europe (EPICE) very preterm birth cohort. International Journal of Epidemiology. 2020;49:372–86. doi: 10.1093/ije/dyz270

29 Gupta S, O’Connor H, Juszczak E, et al. Two-year outcomes after selective early treatment of patent ductus arteriosus with ibuprofen in preterm babies: Follow-up of baby-OSCAR-a randomised controlled trial. EClinicalMedicine. 2025;87:103424. doi: 10.1016/j.eclinm.2025.103424

30 Cohen J. Statistical power analysis for the behavioral sciences. 2nd Edition. New York: Routledge 1988.

31 Baghurst PA, McMichael AJ, Wigg NR, et al. Environmental exposure to lead and children’s intelligence at the age of seven years. The port pirie cohort study. The New England Journal of Medicine. 1992;327:1279–84. doi: 10.1056/NEJM199210293271805

32 Walter T, De Andraca I, Chadud P, et al. Iron deficiency anemia: Adverse effects on infant psychomotor development. Pediatrics. 1989;84:7–17.

33 Swertz M, Enckevort E van, Oliveira JL, et al. Towards an interoperable ecosystem of research cohort and real-world data catalogues enabling multi-center studies. Yearbook of Medical Informatics. 2022;31:262–72. doi: 10.1055/s-0042-1742522

34 Haute Autorité de Santé – Communication and Information Department. METHODOLOGICAL GUIDANCE: Choices in methods for economic evaluation – HAS. Saint-Denis la Plaine, France: Haute Autorité de Santé 2020.

35 Article 30. LOI n 2016-1321 du 7 octobre 2016 pour une république numérique (1). Journal Officiel de la République Française. 2016; 8th October.

